# DROP-DEEP: Dimensionality Reduction for Polygenic Risk Score Using Deep Learning Approach

**DOI:** 10.1101/2024.05.01.24306609

**Authors:** Hadasa Kaufman, Yarden Hochenberg, Michal Linial, Nadav Rappoport

## Abstract

**Motivation:** Advances in sequencing technologies have enabled the early detection of genetic diseases and the development of personalized medicine. However, the variance explained by genetic variations is typically small compared to the heritability estimates. Consequently, there is a pressing need to develop enhanced polygenic risk score (PRS) prediction models. We seek an approach that transcends the limitations of the routinely used additive model for PRS.

**Results:** Here we present DROP-DEEP, a novel method for calculating PRS that enhances the explanation of the heritability variance of complex traits by incorporating high-dimensional genetic interactions. The first stage of DROP-DEEP employs an unsupervised approach to reduce dimensionality, while the second stage involves training a prediction model using a supervised machine-learning algorithm. Notably, the first stage of training is phenotype-agnostic. Thus, while it is computationally intensive, it is performed only once. Its output can serve as input for predicting any chosen trait or disease. We evaluated the efficacy of the DROP-DEEP dimensionality reduction models using principal component analysis (PCA) and deep neural networks (DNN). All models were trained using the UK Biobank (UKB) dataset with over 340,000 subjects and a set of approximately 460,000 single nucleotide variants (SNVs) across the genome. The results of DROP-DEEP, which was established for patients diagnosed with hypertension, outperformed other approaches. We extended the analysis to include an additional five binary and continuous phenotypes, each repeated five times for reproducibility assessment. For each phenotype, DROP-DEEP results were compared to commonly used PRS methodologies, and the performance of all models was discussed.

**Conclusion:** Our approach overcomes the need for variable selection while maintaining computational feasibility. We conclude that the DROP-DEEP approach exhibits significant advantages compared to commonly used PRS methods and can be used efficiently for hundreds of genetic traits.

**Availability and Implementation:** All the codes and the trained dimensionality reduction models are available at: https://github.com/HadasaK1/DROP-DEEP.

## 1. Introduction

The enrichment of Genome-wide association studies (GWAS)^1^, that attempt to identify genetic loci associated with a complex trait by ranking the loci, leaded to new types of models that predict polygenic score (PGS). In the context of disease susceptibility, it is called polygenic risk scores (PRS), also known as polygenic index (PGI). We will refer to all instances as PRS for simplicity. PRS providing an estimate of trait likelihood solely based on genetics and serves as a predictive tool for that trait. The most widely tested markers for this analysis are single nucleotide polymorphisms (SNPs), which are genetic variations that show natural variation in the population^1,2^.

The common approach for PRS aggregates information from SNPs across the genome, by the weighted sum of the trait-associated alleles, into a single score that can be used as a trait value or disease risk prediction for a subject^3–8^. It has been found that the genetic variation explained by trait-associated SNPs does not match the expectations from independent twin studies or classical family studies. Only a small fraction of the genetic risk can be explained by the identified significant SNPs^9^, an discrepancy that refers to the”missing heritability” problem^10^. For example, more than 80% of the variation within a population is attributable to additive genetic factors for height. Still, previous studies were able to find only a small fraction of this variation and the SNP additivity model explained a substantial fraction only in European population^11^.

The conceptual imitation of standard PRS is the assumption of independence between SNPs. Trait-associated SNPs are mostly discovered using single-locus analysis, where variants are evaluated individually for association with phenotypes. In this type of analysis, factors with interaction effects but no independent effects will not be included in the PRS calculation^12,13^. For improving the fraction of explained heritable variance for complex disease, gene to gene (GxG) interactions techniques were developed ^12,13,17^. Unfortunately, in cases that the number of genetic variants are larger than the cohort size, the estimated number of significant parameters introduces to the model faces a genuine combinatorial difficulty^12,14,15^.

Regression-based methods that are widespread data mining methods for detecting GxG interactions in complex traits^14^. For example, the commonly used PLINK tool^16^ utilizes a regression model. Regression-based methods have been criticized due to their inability to deal with a high-dimensional dataset that may contain multi-locus genotype combinations, non-linear relationships, and sparse data. These methods have therefore been largely replaced by more sophisticated machine learning models^14,17^.

Machine learning algorithms, such as artificial neural networks (ANNs) and extreme gradient-boosting (XGBoost), have become practical data mining models in studying associations between genomic data and complex phenotypes because of their ability to learn linear and non-linear gene-gene to phenotype relationships. Thuse models do not rely on most of the prior assumptions that underlie parametric models, and they can capture complex signals from the data and deliver superior predictive accuracy^18^. Previous studies have shown that such models can perform better than linear methods and deliver superior predictive accuracy^19,20^. However, In high-dimensional genomic data, typically there are more parameters (weights) than samples. It may be too computationally demanding when the number of neurons nodes in the network is large and may leads to a problem known as the “curse of dimensionality” with all its implications^21^.

For this reason, many studies have used variable selection techniques in which only subsets of SNPs are used as predictors^18,22,23^, while others have demonstrated that both common and rare SNPs influence traits with minor effects^24^. Previous works have shown that a model using the entire genome has better predictive performance than a model that selects features first. The reason for the lower performance of the latter models is that when SNPs are excluded from the model, a substantial amount of information may be lost, and that cannot be compensated for^25^.

In this study we present DROP-DEEP approach that transcends the limitations of the additive model used in PRS. DROP-DEEP runs dimensionality reduction on the genomic data and then a non-linear prediction algorithm. We start by refining the method for patients diagnosed with hypertension from the UK biobank (UKB) and present the performance with respect to other PRS methods. We then expand the analysis to include additional five binary and continuous phenotypes. For each phenotype, DROP-DEEP results were compared to commonly used PRS methodologies, and the performance of all models was discussed. We finalize by presenting the usability of applying DROP-DEEP for hundreds of human genetic traits and diseases.

## 2. Materials and Methods

### 2.1. Phenotype and Genotype Data

We used the UKB for matching phenotype with genotype for 487,409 samples and 93,095,623 imputed SNPs. We chose six diverse clinical and physiological phenotypes (height, BMI, platelet count, hypertension, type 2 diabetes (T2D), and multiple sclerosis) according to their significant heritability, and high prevalence in the population. Our goal was to examine the quality of the complex trait prediction models. Socio-demographic covariates included age (Data-field 21003), ethnic group (Data-field 22006), sex (Data-field 22001), genotype measurement batch (Data-field 22000), and the assessment center where the patient’s examination was performed (Data-field 54). We also included the first 40 PCA variables (Data-field 22009) calculated by the UK Biobank based on the whole dataset. For platelet count GWAS phenotype, the assessment centers were discarded due to over covariates.

Heritability estimates were extracted from Neale lab heritability browser at https://nealelab.github.io/UKBB_ldsc/index.html.

### 2.2. Pre-processing

Quality control was implemented on participants and SNPs using the PLINK2 tool^16^. SNP quality control was performed by eliminating variants with duplicate IDs, those with missing values (keeping only genotyped SNPs with a dosage of 0, 1, or 2), variants that deviated from Hardy-Weinberg equilibrium (HWE) by p-value < 1e-6, and variants with minor allele frequency (MAF) < 0.001. SNP quality control operations were carried out in the default PLINK2 order. Subject quality control was performed by eliminating family-related subjects, such as Sheppard *et al*. (2021). In addition, we used only subjects of European ancestry to prevent the effects of population structure^2^. The age covariate was min-max normalized to range between zero and one. SNPs were coded as 0, 1, 2 for homozygote for the minor allele, heterozygote, and homozygote for the alternative allele, respectively, assuming additive allele effects. To create 5 repetitions, the samples were randomly split into an 85% training set and a 15% test set. For each repetition, one thousand random samples were taken from the training set to serve as the validation set. Such a large dataset in high dimensions could not be loaded into memory at once. Therefore, for the learning models, we decided to split the data into 1,000 samples each. We used iterative algorithms, meaning the model was updated based on a batch of 1,000 samples in each iteration.

### 2.3. GWAS

For each phenotype separately (5 repetitions), we performed GWAS^1^ using the PLINK2 tool^16^ on the training set. Using this method, trait-associated SNPs are discovered using single locus analysis, where each variant, adjusted to the covariate, is evaluated individually for association with a phenotype. GWAS output was basic for the PRS using PRSice-2, for the PRS using Lassosum, and for the non-linear based SNP-selection models.

### 2.4. PRS Approaches

#### PRSice-2

We performed PRS analyses using the PRSice-2 software^3,26^. In the classic PRS calculation method, only SNPs with a GWAS association p-value below a certain threshold are included in the calculation. The PRSice-2 software searches for a p-value threshold that generates the PRS best fit that maximizes the phenotypic variation. To compute PRS, we used the training set and omitted SNPs with constant dosage. For each SNP, we obtained p-values and coefficients from the GWAS results. Consider SNPs *N* with a p-value smaller than the tested threshold; *x*, a list of the number of effective *x* alleles observed for each SNP, assuming an additive model; and *βi, i* ∈ [ *N* ], a list of the regression coefficients for height and log-odds ratio for hypertension. The PRS is then computed as follows:

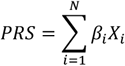

Then, to adjust the PRS to the covariate matrix, we trained a linear regression model for continuous phenotypes and a logistic regression model for binary phenotypes using the Scikit-learn package in Python^27^. The predictors were the PRS obtained from the PRSice-2 software and the covariate matrix.

#### Lassosum

We performed PRS analyses using the Lassosum package^4^. Lassosum is another linear approach to evaluate PRS, which uses Lasso regression. LASSO obtains estimates of *β* (weights in the linear combination) given y (observed values) and *X* (features) by minimizing the objective function:

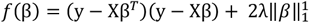

### 2.5. Approaches for reducing number of features

#### Supervised SNP selection

Based on the GWAS results, we calculated for each phenotype (5 times) a threshold for the 10% lowest P-values. The thresholds were 0.096 < p < 0.0173 for all phenotypes. SNPs with lower p-values than the thresholds were used as features for non-linear prediction models (NN and XGB).

#### Unsupervised Dimensionality Reduction

Due to existing resource limits, we trained separate dimensionality reduction models for each of the 22 chromosomes (for the five repetitions, using autosomal chromosomes) for the dimensionality reduction process. Such a large dataset in high dimensions could not be loaded into memory at once. Therefore, we decided to split the data into 1,000 samples each. We used iterative algorithms, meaning the model was updated based on a batch of 1,000 samples in each iteration. We used two models: deep autoencoder and principal component analysis (PCA) ^28^. Other dimensionality reduction alternative methods exist, such as truncated singular value decomposition (SVD)^29^, Lsomap^30^ and t-SNE^31^, and could easily be substituted. However, we chose the two mentioned as they are widely used and can be trained with an iterative process using batch processing. We reduced the dimension of the SNPs to 10% of the input SNP number in each chromosome, a percentage that was adjusted to the computational power available and allowed the later union of the variables from all the chromosomes for the prediction model. For the autoencoder, this was achieved by setting the number of neurons in the encoding layer to 10% of the input SNP number. For PCA, we used the first 10% of the principal components.

The dimensionality reduction phase is independent of phenotype, i.e., training does not require labels, and its output can be used as an input for any type of prediction model and any chosen trait or disease. The dimensionality reduction models were evaluated using the validation set according to the coefficient of determination metrics (see evaluation metrics section).

We trained a fully connected autoencoder model with the TensorFlow Python package^32^. Each chromosome’s autoencoder was designed as an encoder with two hidden layers, with the second layer being the feature compression layer (bottleneck) and a decoder with one hidden layer. We chose to add dropout layers to prevent overfitting and because this technique is known for its ability to efficiently combine many different neural network architectures^33^. Each hidden layer had a dropout layer with a rate of 0.1, except for the bottleneck. The number of neurons in each hidden layer was 20%, 10%, and 20% of the input SNP number, respectively. We performed hyperparameter tuning using the validation set for the activation functions and for the learning rate, where the best configuration was chosen according to the RMSE metric. As hyperparameter tuning is a resource-intensive task, which takes long run times, we performed the tuning on chromosome 22 and used optimal hyperparameters for the other chromosomes. We trained our model using the Adam Optimizer^34^ and the mean squared error (MSE) loss function with a batch size of 250 over 500 epochs. The chosen activation functions were PReLU^35^ for the hidden layers and linear for the output layer, and the best learning rate was 1e-5.

The second method for dimensionality reduction we used was PCA, implemented by the Scikit-learn package in Python^27^. We projected the SNP data of each individual on each chromosome using the first 10% of the principal components (PCs). The data after dimensionality reduction was min-max normalized to be between zero and one.

### 2.6. Phenotype Prediction

Each prediction model was trained (5 repetitions) on the 10% SNP selection data, on the 10% layer from the autoencoder, and on the 10% first PCs. We concatenated the new representations of each chromosome and adjusted them to the covariate matrix. The combined datasets were used as inputs for the phenotype prediction model. We used two models for prediction: deep neural network (DNN)^36^ and XGBoost (XGB)^37^. Each model had its hyperparameters tuned, and the best configuration (Supplementary Table 1) was chosen using the validation set according to the RMSE metric for height and the log-loss metric for hypertension. The models were then evaluated on the test set using different evaluation metrics. The training run times of the different models are shown in Table 2 (see resources section for information about the resources consumed).

**Table 1.** Prediction results on hypertension using the test set. The results represent here are the averages for 5 repetitions.

| Method<sup>a</sup> | ROC AUC<sup>b</sup> | Log Loss | Average Precision |
| --- | --- | --- | --- |
| PRSice-2 | 0.5782 | 0.5950 | 0.2745 |
| lassosum | 0.5997 | 0.5591 | 0.2934 |
| SNP selection + XGB | 0.6905 | 0.4992 | 0.3553 |
| SNP selection + DNN | 0.6455 | 0.5782 | 0.3335 |
| PCA + XGB | 0.6812 | 0.5148 | 0.3445 |
| PCA + DNN | **0.7104** | **0.4879** | **0.3913** |
| AuEn + XGB | 0.6814 | 0.5029 | 0.3433 |
| AuEn + DNN | 0.7038 | 0.4887 | 0.3849 |

<sup>a</sup>AuEn stands for Autoencoder. <sup>b</sup>ROC AUC, Area under the ROC Curve

**Table 2.**
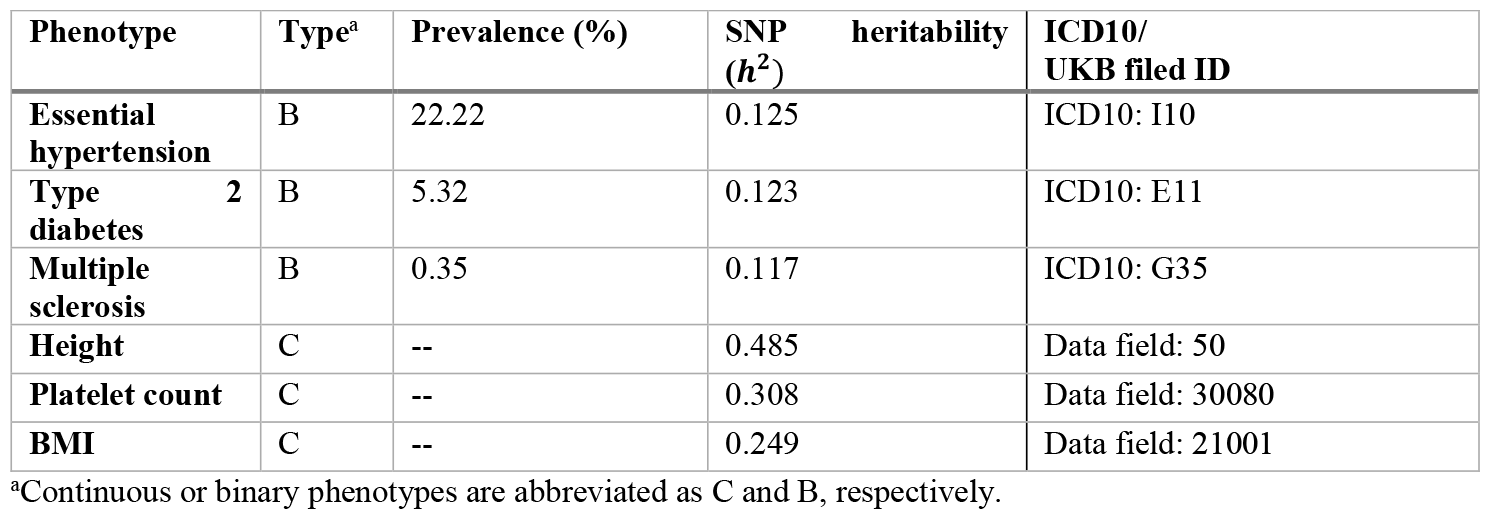
Phenotypes that were used to test the DROP-DEEP approach.

We trained a fully connected DNN model with TensorFlow^32^. The network was designed with two hidden layers, and each hidden layer had a dropout layer with a rate of 0.1. The number of neurons in each hidden layer was 20% and 10% of the number of input variables, respectively. We tuned the hidden layers’ activation function for each model (**Table 1**), and we chose a linear output layer activation function for height and a sigmoid for hypertension. We trained our model using the Adam optimizer^34^ with a batch size of 50 over 80 epochs. The learning rate used was 1e-7, and the loss function was MSE for continuous phenotypes and binary-cross-entropy for binary phenotypes. We trained an XGB model with the XGBoost Python package for the second prediction model. XGB can be used for regression (for height) and classification (for hypertension). We trained the model in batch processing using the “xgb_model” parameter, which allows training continuation. For each batch of 1,000 samples, the model built one gradient-boosted tree, using a 0.2 subsample ratio of columns and a 0.01 minimum loss reduction required to make a further partition on a leaf node. We tuned the maximal depth of the tree for each model separately (**Supplementary Table S1**), and we trained our model for 20 epochs for binary phenotype prediction and 40 epochs for continuous phenotypes prediction until convergence. The learning rate used was 1e-3, and the evaluation metric was RMSE for continuous phenotypes and binary-cross-entropy for binary phenotypes.

### 2.7. Evaluation Metrics

We compared the correct SNP values and the reconstructed SNP values obtained from the decoder models to evaluate and compare the conserved information of dimensionality reduction models. We did this using the coefficient of determination (R2) metric that was calculated using the weighted variance aggregation implemented by the Scikit-learn package in Python^27^.

For evaluating and comparing the different algorithms for complex trait prediction, we chose the metrics according to the phenotype type. For quantitative phenotypes, we used R2 and root mean square error (RMSE). Consider *N* samples, a list of correct values *yi*, and a list of predicted values obtained from the estimated model *yi, i* ∈ [*N*]; the RMSE is calculated as follows:

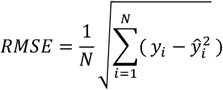

R2 is calculated as follows:

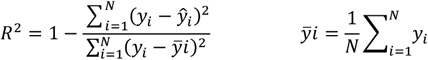

For binary phenotypes with unequal distribution between the two classes, we used the Area Under the Receiver Operating Characteristics (ROC AUC), Average Precision metric, and Cross Entropy Loss (log-loss) metrics. Consider N samples, a list of estimated probabilities obtained from the estimation model *pi*, and a list of correct values *yi, i* ∈ [*N*]; the log-loss is calculated as follows:

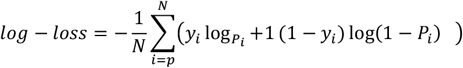

Considering t1…*tn*] range of thresholds, the Average Precision is calculated as follows:

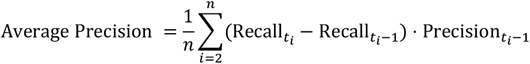

where *Precision* is the proportion of true positive samples in the total predicted positive observations, and *Recall* is the portion of true positive samples in the total predicted negative observations.

## 3. Results

### 3.1. Overview of the DROP-DEEP approach

DROP-DEEP combines dimensional reduction of genotype data, and deep learning (DL) models to predict PRS of continuous and binary phenotypes. In a nutshell, we first carried out quality control (QC) and continue our analysis with 341,985 genotypes with 467,429 SNPs from the UKB. Each feature was presented as accepted in the field with 0, 1 and 2 associated with the absence of the SNP, its present in one or both alleles, respectively. Then, we used unsupervised PCA to reduce the number of parameters to 10% of the original number of features. We scaled the 10% PCs min-max scale, and then we appended all chromosomes to the complete dataset and trained neural networks (NNs). DROP-DEEP was built while comparing to the autoencoder in the dimension reduction phase and the tree-based model in the learning phase and was chosen as the most efficient model for its reduced runtime and the good predictive performance. This model was tested with six different phenotypes (with five repetitions each) that were rigorously compered to routine approaches. We show that significant advantages are associated with binary phenotypes of complex diseases **(Fig. 1)**.

**Fig. 1.**
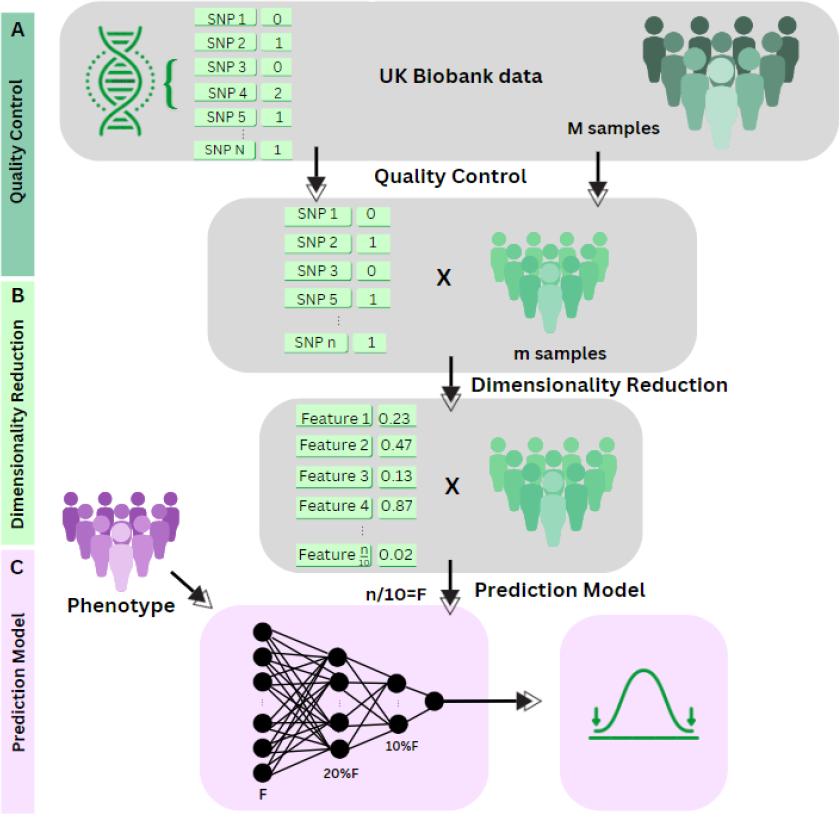
An overview of DROP-DEEP methodology. **(A)** The first stage includes the quality control and choosing SNPs according to MAF <0.001, HWE (p-value > 1E-6) and removing the SNPs with missing values. Samples quality control including filtering out samples from non-European origin and keeping only a representative from each kindship (i.e., SNP decreased from N to n, and samples from M to m). **(B)** Unsupervised dimensionality reduction using PCA reduces the number of features to 10% of the n SNPs while keeping maximum variance included in the original data. Therefore, we overcome the need for predetermine feature selection. **(C)** The last stage is phenotype dependent. The reduced new features (F) are used as input for training NN, and a PRS score is calculated.

### 3.2 Comparing alternative PRS models

We demonstrate our method on patients from UKB that were diagnosed with Essential hypertension (ICD-10: I10). To maximize the performance of DROP-DEEP, we ran DROP-DEEP with two dimensionality reduction methods and two prediction methods. To assess the method, we compared the performances to the two most routine common approaches of PRS: Lassosum and PRSice-2. In addition, we compare the results to a standard GWAS based on SNPs selection and then prediction with learning model (**Fig. 2**).

**Fig. 2.**
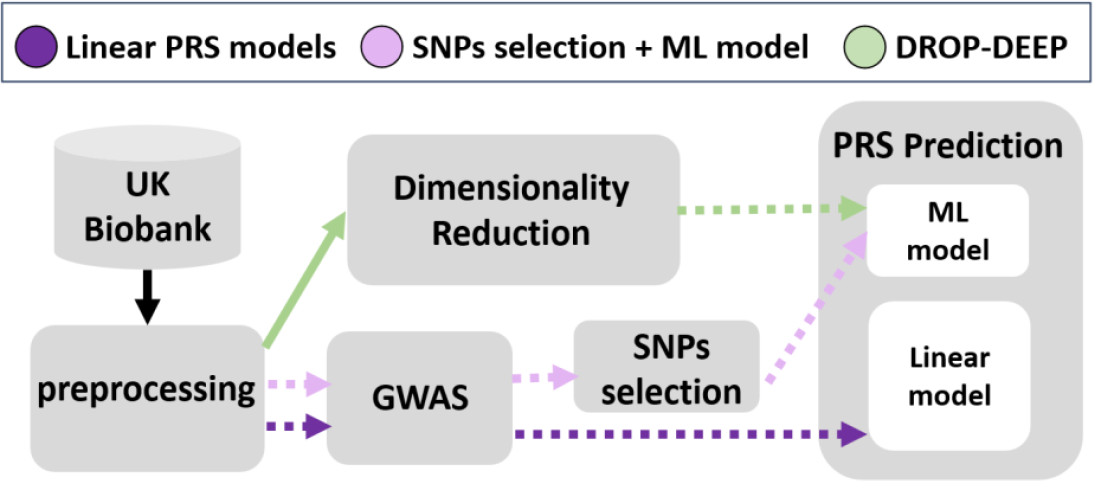
Schematic overview of the study framework. DROP-DEEP approach is indicated with a green color while common PRS approaches are marked with in light purple, and the classical GWAS-based SNPs selection, followed by prediction is indicated by dark purple color (for further details see Supplementary Fig. S1). Dotted arrows represent phenotype-dependent analysis and continuous arrows represent phenotype-independent analysis. The purple pathways are phenotype-dependent from the beginning while the DROP-DEEP approach became phonotype dependent only at the ML model training step.

### 3.3. Pre-processing and feature reduction

Altogether, after applying a quality control filter, there were 341,985 samples and 467,429 SNPs left for analysis. These samples were partitioned to 289,688 samples for the training set, 51,297 samples for the test set and validation set includes 1,000 samples. For the showcase of hypertension diagnosis, we collected 75,984 cases and 266,001 controls. The number of cases and controls in each repetition training and test set can be found in Supplementary **file S1**.

The dimensionality reduction results were evaluated using the validation set for each chromosome and each dimensionality reduction model. The weighted average *R*^2^ for the autoencoder achieved 0.5745, and for the PCA it achieved 0.6642, where each chromosome’s *R*^2^ was weighted according to the number of SNPs (see Supplementary **Table S2**). The autoencoder’s *R*^2^ yielded lower results than the PCAs in all chromosomes.

As shown in **Fig. 3A**, the average training time for the autoencoder for 100 epochs as well as the GWAS running time, was much longer than the total time needed for training the PCA. The autoencoder was trained up to 500 epochs that have been shown minor changes in the log loss values. The accumulated variance of 10% of the PCs was ∼0.7 for all chromosomes (**Fig. 3B**).

**Fig. 3.**
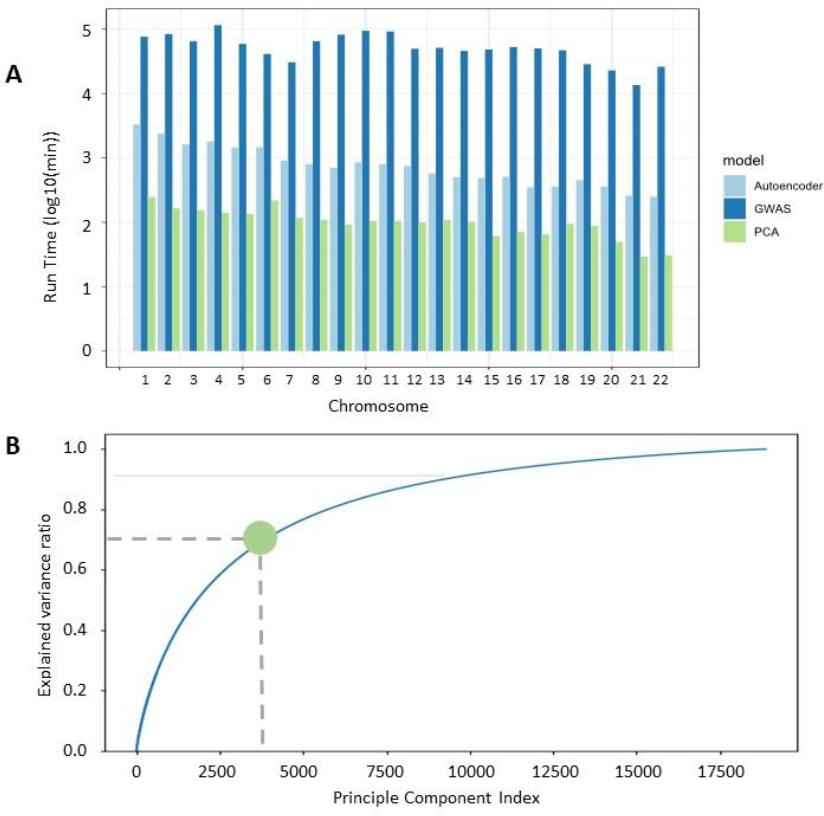
Metrics for dimension reduction methods. All the metrics display are from repetition 1. **(A)** A compression between the running time among methods of feature reduction. The running time displayed for the autoencoder model is the average of the running time of each 100 epochs (out of total of 500). The running time for GWAS is the average time for hypertension GWAS. **(B)** The accumulated explained variance as function of the number of PCs is illustrated for chromosome 1. The green circle represents 10% PCs.

### 3.4. Hypertension predictive performance

The total number of features in the prediction model input matrix was 42,870, including 42,742 variables from the dimension reduction process unified from all 22 autosomal chromosomes. In addition, there are 128 variables from the covariate matrix that included age, sex, 105 dummy variables of the batches used for the genotype measurement, and 21 dummy variables of UKB assessment center. The results for the PRS models for all the approaches is summarized in **Table 1**.

**Table 1** shows that the non-linear methods outperform the common PRS approaches (PRSice-2 and lassosum). Among all the approaches based on the reduction of the number of features followed by a learning model for prediction, PCA with DNN provided the best results for all measures, with ROC AUC of 0.7104, the lowest log loss (0.4879) and the highest average Precision (0.3913). Notably, the autoencoder with DNN shows good results but with much longer running time.

### 3.5. Generalized predictions

In order to examine whether DROP-DEEP improves the predictions for other phenotypes, we chose five additional phenotypes from the UKB cohort. We included binary phenotypes of complex diseases with a broad prevalence range (22.2% to 0.35%). In addition, we analyzed continuous traits with relatively high heritability. The prevalence was calculated from the UKB data that was used for this study. The list of phenotypes is shown in **Table 2**.

For hypertension and type 2 diabetes DROP-DEEP displayed the best results (**Fig. 4A**), while in disease of multiple sclerosis (with very low prevalence, **Table 2**), there was no significant difference than PRSice-2 which displayed the best results. For height and platelet count, the SNP selection with NN displayed the best results (**Fig. 4A**).

**Fig. 4.**
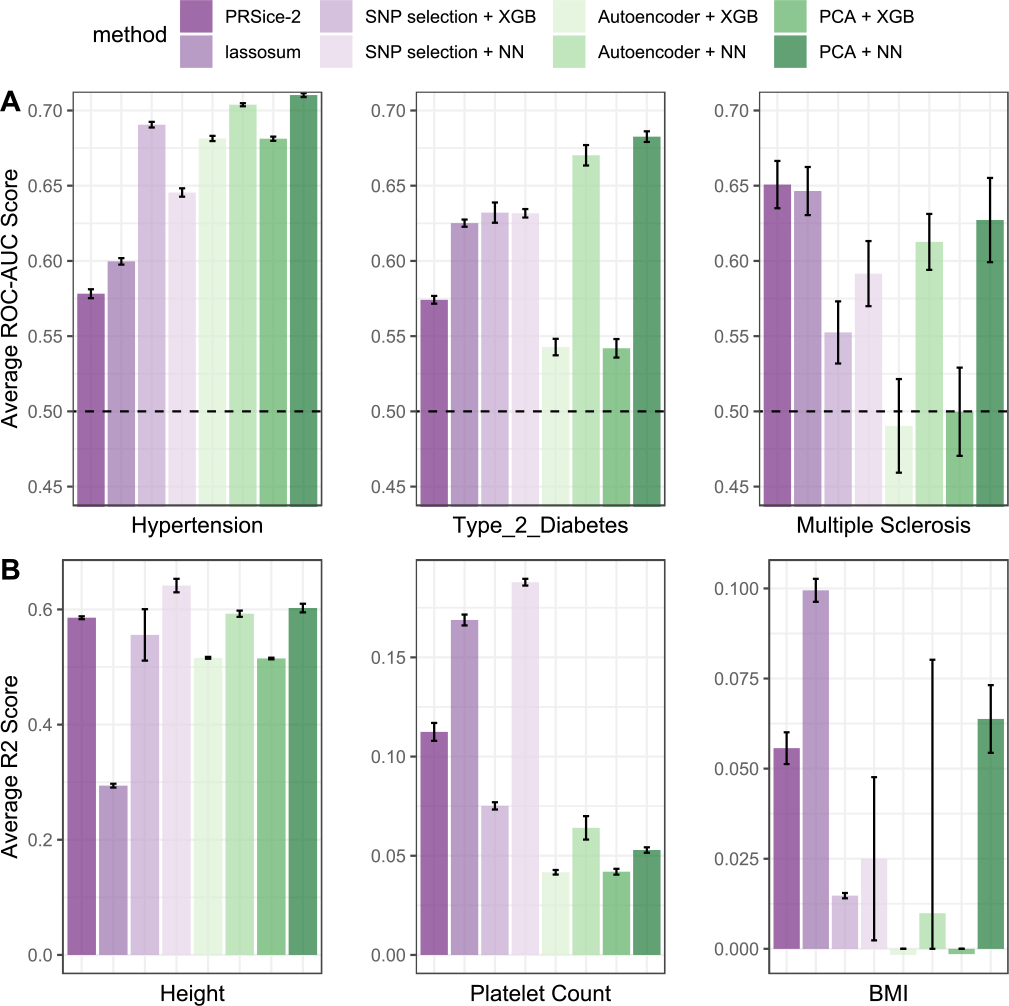
Comparison between different PRS approaches discussed in this study. The scores shown are for validation set of 1000 samples. (A) For the binary phenotypes ROC-AUC score represent the prediction quality. Each column in the data displays the average and standard deviation of 5 repetitions (for further details see Supplementary **Table S3**). **(B)** Comparison between the R^2^ score of the continuous phenotypes (for further details see Supplementary **Table S4**). Note the different scale for the R2 value. All the metrics for each of the phenotypes are available in the supplementary data. Note the different scale of the y-axis.

For 5 of the 6 selected phenotypes, we observed high consistency between repeated runs), an exception is the BMI (body mass index) which associated with noisy *R*^2^ results for the models based on NN prediction following the SNP selection or the autoencoder. For 5 of the 6 phenotypes, the model of PCA with NN surpassed all other 3 paths in the unsupervised dimensionality reduction with the learning model approach. For almost all listed phenotypes, the autoencoder with NN displayed similar but slightly lower results with a much longer run time.

The XGB models were consistently significantly worse than the NN models after dimensionality reduction, but after SNP selection it showed close or often even improved results relative to NN model, only for the binary phenotypes. Interestingly, we observed differences in the trends of the linear PRS model results and the learning-based PRS models. For example, for both PRSice-2 and lassosum, the ROC AUC score of multiple sclerosis was higher than the hypertension and type 2 diabetes ROC AUC score, while the heritability estimate was lower for multiple sclerosis. The DROP-DEEP approach has shown a positive correlation between the ROC AUC score the heritability (0.56) while an inverse correlation was detected for PRSice-2 and lassosum (−0.77 and -0.34 respectively). In the continuous phenotypes (Table 2), both the DROP-DEEP approach and the linear approach displayed very high correlation (>0.95), but it is noticeable that the differences in the lassosum performances between the less heritable phenotypes and the height phenotype are significantly lower than the differences between the phenotypes in all the learning-based models. Importantly, the reproducibility of the results (upon repeated runs) was mostly phenotype-dependent rather than methodology dependent.

## 4. Discussion

The main notion of PRS claims that the accumulation of multiple risk alleles contributes to an individual’s genetic liability for phenotypes such as complex diseases. The PRS is a quantitative measure representing an individual’s genetic susceptibility to a specific trait. It is calculated by summing risk values assigned to genetic variants associated with a phenotype of interest. These variants are weighted by their effect sizes, typically derived from GWAS^1^. Those PRS approaches assume additive effects, neglecting potential interactions between variants. However, it is clear that epistatic effects present and expected to impact human traits^38^. Also approaches proposed in recent years, based on learning models and enabling the learning of more complex interactions, are founded on GWAS-based feature selection before. For all these methods, the caveat is that PRSs constructed from pre-selected genetic variants might not capture the entirety of genetic liability. Moreover, rare variants that often fails to reach statistical significance, are not covered or identified by GWAS and therefore are excluded from PRS calculation.

Selection of human traits of different nature stems from the known difference for the impact of additive genetics for the phenotype. For example, height shows consistent estimates of heritability between twin studies, family studies and SNP estimates. On the other hand, the *h*^2^ estimates for BMI are much higher from twin studies relative to family studies, suggesting non-additive effects to be important for BMI but less so for height.

Given the above limitations, we present DROP-DEEP, a novel PRS calculation method that enhances the ability to predict complex traits based on heritable variation. DROP-DEEP takes genetic interactions into account and is not based on GWAS or prior knowledge. Therefore, can account for variants that would be omitted by other approaches. Importantly, the dimensionality reduction step is independent of the phenotype, thus it can be performed once and used for any desirable phenotype, clinical or physiological, with only the predication model training step needing to be redone for each phenotype. DROP-DEEP showed improved predictive ability with high consistency for the binary phenotypes and reduced runtime relative to learning-based alternatives PRS approaches.

We compared our method to established methods in terms of (i) quality of results, (ii) running times, and (iii) stability. Considering the results, we recommend DROP-DEEP as the most efficient method for binary phenotypes (**Fig. 4A**), both in terms of running times and in terms of the quality of the results. For continuous phenotypes (**Fig. 4B**), running times of GWAS-based SNP selection are acceptable and result in better results. Thus, is recommended as a protocol of choice for such cases. At the same time, for those who calculate PRS for a large number of phenotypes, binary and continuous, DROP-DEEP gives a result not far from the maximum for all the phenotypes we tested and can significantly save running time by reducing the dimension once for all the phenotypes, and no GWAS is required for each individual phenotype. Although the autoencoder should be able in principle to capture non-linear relationships between SNPs, it was consistently significantly longer than PCA and did not improve the results relative to it, so we do not recommend using it in such a case.

We acknowledge certain limitations in this study. First, our model was trained and evaluated only on the European population from UKB. Thus, the transferability and adaptations of the predicting model across ethnic groups may be limited. Second, we tested the model on selected six phenotypes that represent a range of heritability estimate and prevalence. However, there are hundreds of phenotypes that are available in UKB and other resources that are of great interest to the biomedical community. We tuned the PRS-based recommendations according to the tested phenotype. An extension of the comparison to a wider range and types of phenotypes could substantiate our recommendations. This is of special importance for gaining more stable and evidence-based recommendation to PRS algorithm that can apply also for low prevalence phenotypes. Moreover, we conducted our study on a subset of all SNPs (avoiding missing reads) for the sake of saving computational burden. In order to robustly recommend optimal predictions from this study, it is necessary to expand the number and representation of the SNPs to the full range (approximately 10 million SNPs) of high quality SNPs that represent the population genetic variability. Lastly, useing of external independent dataset for validation is critical for generalization purposes and this is of an even more crucial in the case of non-linear interaction between genetic variants^38^.

In conclusion, DROP-DEEP is a new approach to PRS based on dimensionality reduction of human SNPs. We recommend using this approach for binary phenotypes, where it shows improvement over all other approaches across multiple phenotypes. The new approach can be used to develop personalized medicine strategies and improve health services.

## Data Availability

All data in the study is based on the UKB

## Supplementary Information

Supplementary data are available in the Supplementary_Data.docx file. The data underlying this study were provided by the UKB under license (Application ID 26664).

